# Multi-sector stakeholder consensus on tackling the complex health and social needs of the growing population of people leaving prison in older age

**DOI:** 10.1101/2023.04.27.23289227

**Authors:** Ye In (Jane) Hwang, Adrienne Withall, Stephen Hampton, Phillip Snoyman, Katrina Forsyth, Tony Butler

## Abstract

As populations age globally, cooperation across multi-sector stakeholders is increasingly important to service older persons, particularly those with high and complex health and social needs. One such population is older people entering society after a period of incarceration in prison. The ‘ageing epidemic’ in prisons worldwide has caught the attention of researchers, governments and community organisations, who identify challenges in servicing this group as they re-enter the community. Challenges lie across multiple sectors, with inadequate support leading to dire consequences for public health, social welfare and recidivism. This is the first study to bring together multi-sector stakeholders from Australia to form recommendations for improving health and social outcomes for older people re-entering community after imprisonment. A modified nominal group technique was used to produce recommendations from N=15 key stakeholders across prison health, corrections, research, advocacy, aged care, community services, via online workshops. The importance and priority of these recommendations was validated by a broader sample of N=44 stakeholders, using an online survey. Thirty-six recommendations for improving outcomes for this population were strongly supported. These recommendations entail two important findings about this population: (1) They are a high-needs, unique, and underserved group at risk of significant health and social inequity in the community, (2) Multi-sector stakeholder cooperation will be crucial to service this growing group.

## Introduction

Worldwide, the number of older people in prisons is growing (Ginnivan et al., 2021; Merkt et al., 2020). This ‘graying’ of prisoner populations worldwide emerged in literature in the early 2000s, with recognition of the phenomenon and its implications across multiple countries (Ahalt et al., 2013; Fazel et al., 2001; Ginnivan et al., 2021; Reimer, 2008). It has been referred to as a crisis, in recognition of the complex and multifaceted challenges it presents across justice, health and welfare sectors (Maschi et al., 2013; Scaggs & Bales, 2017). It is driven by factors including general population ageing, longer sentences, convictions for historical offences in later life, and trajectories of repeated imprisonment in certain high-risk groups (Howard & Corben, 2019; Luallen & Cutler, 2017; Roth, 2014; Scaggs & Bales, 2015). In Australia, the total prisoner population rose by 47% from 2009-2019, but the number of prisoners aged 50+ and 65+ rose by 81%, and 142% (Australian Bureau of Statistics, 2009, 2019). This rate of growth exceeds that of the general community during the same period (25% and 81%). Up to 1 in 4 people in prisons across the world are now older, and this proportion continues to grow (Australian Institute of Health and Welfare, 2019; Merkt et al., 2020).

Most people in prison will be released back into the community, and successful reintegration of this group is a challenge with older people in prison having poorer physical and mental health, and higher unmet health and social needs compared to both younger people in prison, and older people in the community (Australian Institute of Health and Welfare, 2019; Lee et al., 2019; Solares et al., 2020; Stevens et al., 2018). This challenge is exacerbated by prison systems that have been traditionally focused on security rather than care, and the stereotypical ‘young’ inmate (Davies, 2011; Hwang et al., 2021). These multiple vulnerabilities has resulted in consensus that in the prison context, people over 50 should be considered ‘older’ (Merkt et al., 2020). Resultantly, this group leave prison with the complex health and social service needs that are already associated with justice-involved populations, but further compounded by premature ageing, as well as the social challenges and stigma associated with incarceration.

One review concluded that older people leaving prison suffer disproportionate challenges due to factors including: reduced social supports in the community, heightened health and mobility support needs, stigma, and difficulty adjusting psychologically following a life of institutionalisation (Davies, 2011). More recent literature confirms that these issues remain pertinent. For example, one study of prison leavers (N=101, Age 55+) in the U.S. found 46% of their sample had visited an emergency department within 6 months of release, with 21% having visited more than once (Humphreys et al., 2018). An Australian study reported that over half of its sample of released prisoners aged 45+ (N=1,853) experienced discontinuity of their mental health care needs after release (Sodhi-Berry et al., 2015). Qualitative work with this group and those involved in their care also highlights that reintegration difficulties for this group are manifold, including difficulties dealing with parole requirements, stigma, homelessness, job discrimination, technology use, essential life skills, seeking support services, rebuilding social connections, and managing multiple health conditions (Hwang et al., 2022; Jimenez et al., 2021; Lares & Montgomery, 2020; Wyse, 2018). People who have left prison in older age thus represent a highly medically and socially vulnerable population in the community, whose unmet needs carry costly consequences not just for the individual’s health and wellbeing, but also for health equity, social services, and recidivism.

In Australia, a growing body of literature recognises the rapid ageing of the prisoner population with qualitative studies, economic analyses, examination of offending data and commentaries conducted by academics and various government agencies all acknowledging this issue (Australian Institute of Health and Welfare, 2019; Ginnivan et al., 2021; Hagos et al., 2021; Howard & Corben, 2019; Hwang et al., 2021; Inspector of Custodial Services NSW, 2015; Office of the Inspector of Custodial Services WA, 2021; Simpson et al., 2017; Trotter & Baidawi, 2015). Whilst mostly focused on prisoner population ageing or prisoner health management in general, all emphasise the importance of appropriate post-release support for older prison leavers. One qualitative study with correctional staff (N=32) in Australia specifically focused on investigating the specific barriers and enablers to post-release reintegration in older prison leavers, and identified that the challenges exist at not only the personal, but also the social, economic and organisational levels (Hagos, 2021).

Overall, the literature from both Australia and worldwide indicate that the concerted effort of multiple stakeholders across health, justice, research and social services both in prisons and in the community is needed to ensure successful reintegration of this group into the community (Hagos, 2021; Metzger et al., 2017; Schwartz et al., 2020). Metzger and colleagues’ 5-step COJENT framework (Criminal Justice Involved Older Adults in Need of Treatment) Initiative provides a useful framework to “determine the needs of criminal justice-involved older adults, assess the community’s relevant resources, identify knowledge and resource gaps, and to use this information to develop a stakeholder-driven action plan to address their needs” (Metzger et al., 2017, p. 2). The 5 steps include: (i) identifying multi-stakeholder perceptions of key issues, (ii) conducting community-based needs assessments, (iii) implementing quick-response interventions, (iv) holding public forums to engage stakeholders to prepare for action (v) consolidating the evidence and engaging “champions” to collaboratively develop and deliver an action plan.

This study aimed to bring together multiple stakeholders to form consensus on recommendations for improving health and social service delivery for older people leaving prison in Australia to gain novel insights into key areas of need for this growing population. It also aimed to identify current strengths, resources and opportunities available to these stakeholders to begin working towards solutions. The study was designed in broad alignment with Step 1 of the COJENT framework and gathered useful information for informing Steps 2 and 3.

## Method

### Design

A modified version of the nominal group technique (McMillan et al., 2014, 2016) was used. The nominal group technique is a flexible, consensus-building method that allows for collaborative agreement to be reached whilst empowering each participant to contribute and have their ideas considered by others (McMillan et al., 2014). The original design by Delbecq et al (1975) involves four stages: *Silent Generation of Ideas* done individually, *Round Robin* sharing ideas as a group, *Clarification* of ideas, and *Voting* for ideas.

In this study, we introduced several variations to build on the strengths of nominal group technique and to enhance utility and validity of the findings (Figure 1). Dashed boxes indicate the four stages of a traditional nominal group technique. The key variations were as follows. First, we improved participants’ ability to make better-informed recommendations by providing the ideas generation question a week prior to the workshop date, offering a brief presentation on the current state of literature at the beginning of the workshop, and including a ‘warm up’ discussion question. Second, we hosted multiple smaller workshops rather than just one, hence eliciting more in-depth data from the fewer participants in each workshop and reducing the burden on participants to commit to a longer workshop. Finally, in addition to the workshop participants, we recruited a larger sample of the relevant population to complete the voting stage. This allowed a broader and endorsement of the final list of recommendations.

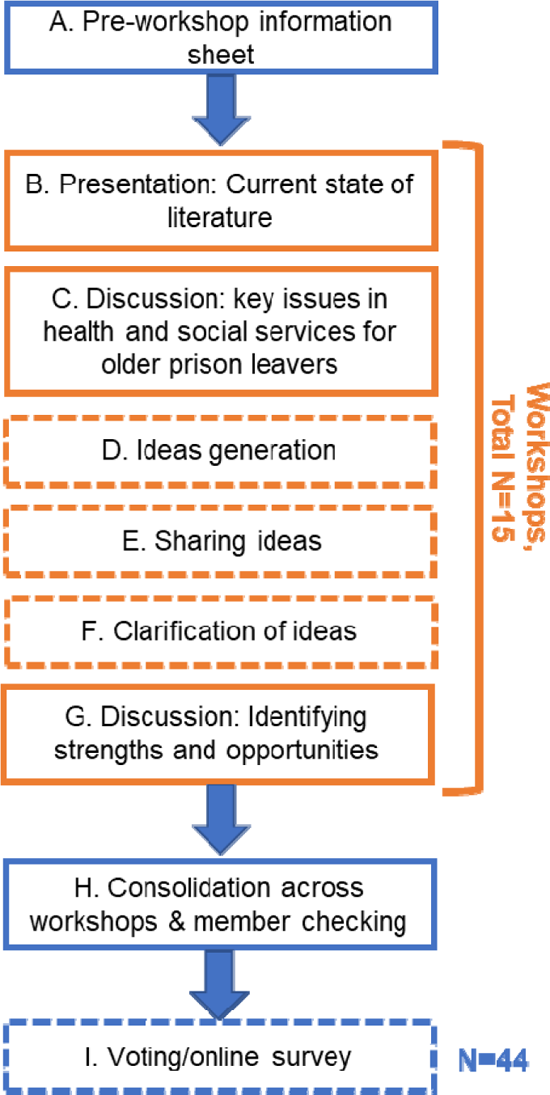

At the end of the workshop was a discussion question for participants to identify any existing strengths, opportunities or resources that they are aware of in their organisations or in the community that may encourage positive change for older prison leavers. The current paper focuses on the development of priorities from these workshops (ie steps D-I in Figure 1). The results of the key issues for older prison leavers, as well as current strengths and opportunities (ie Step C and G in Figure 1) are described elsewhere (Hwang et al., preprint).

### Ethical approvals

This study was granted ethical approval from: The University of New South Wales Human Research Ethics Committee [HC220042], Corrective Services NSW Ethics Committee [D2022/0294030], and the Justice Health and Forensic Mental Health Network of NSW Ethics Committee [G477/22].

### Sampling, recruitment and consent

#### Workshop Participants

Recruitment was targeted at six types of stakeholders: in-prison staff (health and custodial), community corrections, post-release transition support services, aged care providers, advocacy groups (older persons and justice-involved), and research academics. Purposeful critical case sampling with snowball sampling was used to identify a relatively small number of participants with specific expertise from a large pool of potential participants, with roughly equal representation across all the relevant stakeholder groups. The research team contacted key people in each of the organisations across Australia via publicly available details and worked with them to identify suitable participants.

Inclusion criteria:

– At least 12 months professional experience dealing with older prison leavers in Australia
– Age 18+

Exclusion criterion:

– Experience with older prison leavers restricted to more than 5 years ago

Participants were invited to the study via an email invitation, that also included a copy of the Participant Information Statement and Consent Form, and a study flyer. Participants were able to express interest by contacting the research team via email or telephone. Upon expressing interest, the research team conducted an eligibility screen and answered any questions. Consent was collected via an online form.

### Survey Respondents

For the online survey, we aimed to recruit a larger sample of relevant stakeholders to rank the recommendations that were developed during the workshops. After the workshops were complete, the same organisations as above were re-contacted, with a request for them to disseminate information about the ranking activity to their staff broadly.

### Workshops

Workshops were conducted online via Microsoft Teams and ran for 90 minutes. Workshops were mediated by the first author and a research assistant. Video recordings of the workshop were downloaded at the end of each workshop and converted to audio-only files. These audio files were transcribed by the research assistant. Fifty Australian dollars were available to participants as renumeration for their time. However, some participants who were employed by government agencies were unable to accept this payment.

### Consolidation of Recommendations and Member Checking

The workshop mediator and a research assistant worked together to consolidate the recommendations from each workshop. Potentially overlapping ideas were discussed and clarified by re-listening to each of the workshops. Member checking was then conducted by sending the consolidated list of recommendations, with rationale for all newly formed recommendations or those that were merged due to duplication, to the workshop participants via email. Participants were given two weeks to respond to this email with any feedback.

### Online Survey

The final list of recommendations was transferred onto an online survey on Qualtrics and sent back to the workshop participants, as well as the expanded sample. The survey included screening questions regarding relevant expertise/interest in older prison leavers. After this, participants were presented with the list of recommendations and asked to score each on a Likert scale, according to how important they believed the recommendation was (1=not important, 9=extremely important). They were then asked to select the top 7 that they believed were priority for implementation (in order). Participants could enter a draw to win an online gift card 100AUD in value.

### Survey Analysis

The survey was open for three months. The researchers manually reviewed the screening questions to ensure only eligible participants’ responses were included for analysis.

We referred to methods by McMillan and colleagues regarding analysis across multiple groups in nominal group technique (2014). Ranking scores were imported into Microsoft Excel. All items were then ranked according to medians. Medians of 7-9 were defined as ‘strong support’, 4-6 as moderate, and 1-3 as weak. The level of support for each item was assessed by the median score, and the level of agreement within each group of similar stakeholders was assessed using the mean absolute deviation from the median.

Scores for the top 7 priorities for implementation were calculated using a weighted sum approach. Items ranked higher were given a higher weighted score, i.e., (ranked 1^st^ = 7 points, 2^nd^ = 6 points, 3^rd^ = 5 points, 4^th^ = 4, 5^th^ = 3, 6^th^ = 2 points, 7^th^ = 1 point). A final score for each item was calculated based on the sum of these weighted values.

## Results

### Workshops

Workshops were conducted in June 2022, and N=15 participants took part. Whilst we cannot detail the specific experience represented by each of the stakeholders for privacy concerns related to small sample size, the critical case sampling employed ensured that those with strong expertise on the topic were identified to participate in the study. Each participant often noted having numerous years of experience across multiple roles within justice health, corrections and transition support across Australia. The academics all had a combination of publishing papers, acquiring research funding and running research projects on the topic of older incarcerated people in Australia. All stakeholders were from two states: New South Wales and Victoria.

**Table 1.** Nature of participants in consensus workshops

| Stakeholder type | n |
| --- | --- |
| Prison health provider | 3 |
| Corrective services (custody and community) | 3 |
| Post-release transition support services | 4 |
| Advocacy and community groups | 2 |
| Researcher | 3 |

The workshops resulted in a final list of 37 recommendations that could be divided into nine categories (Table 2; also see Appendix 1). A brief description of the categories is provided in Table 2.

**Table 2.**
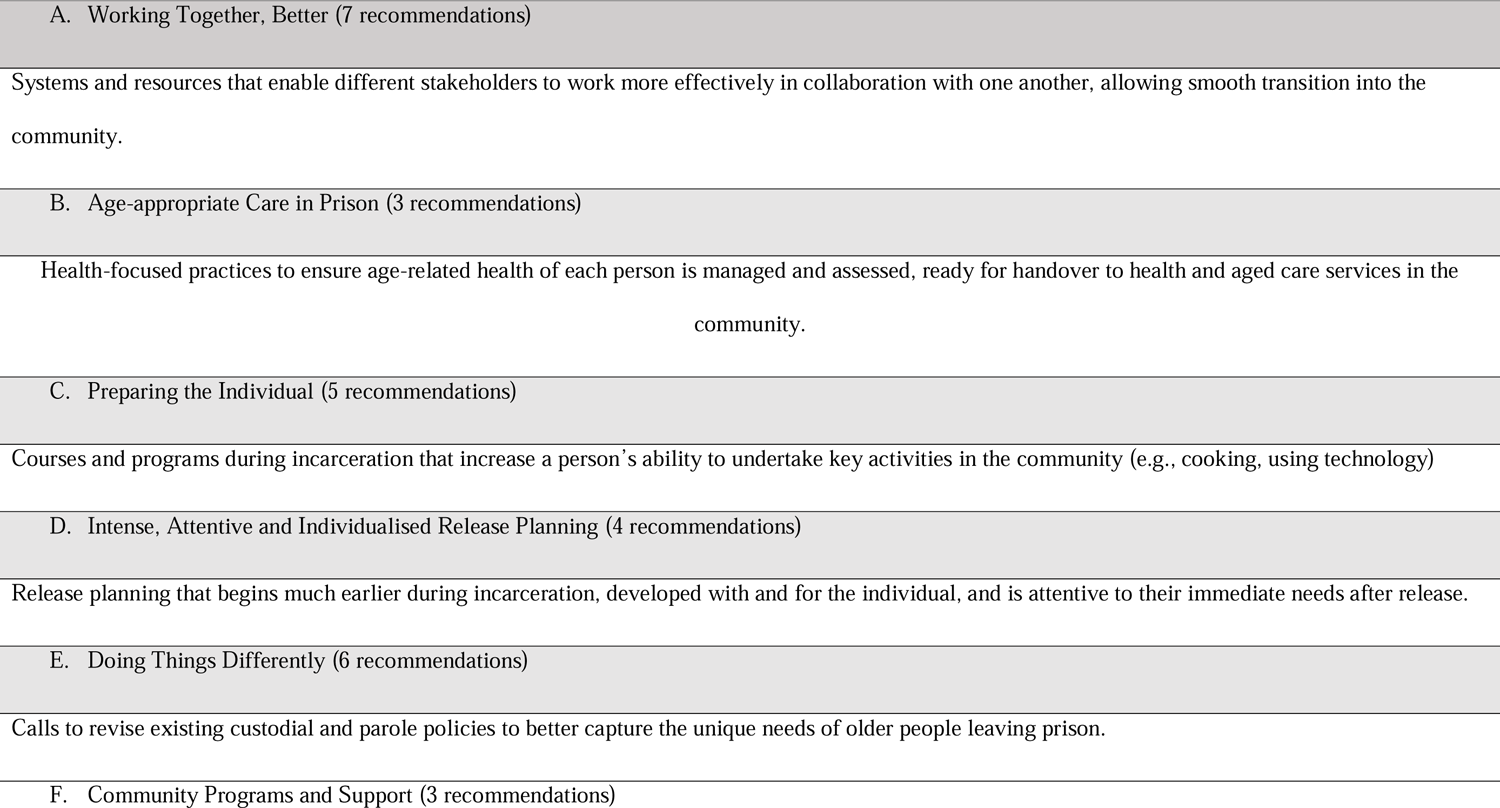
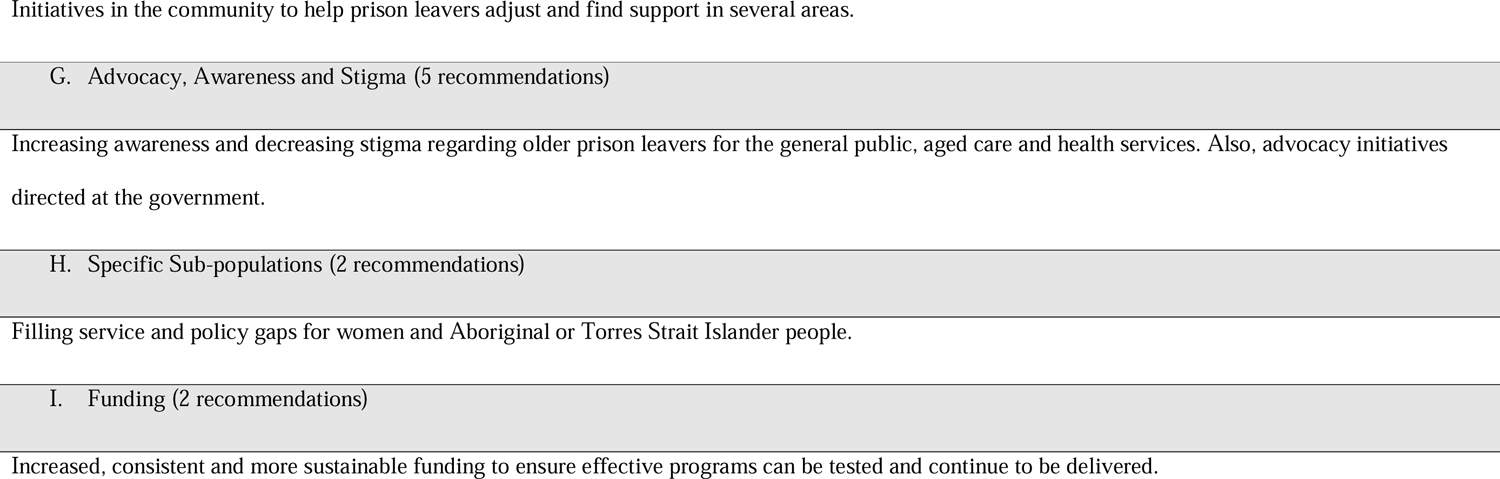
Categories of final recommendations

### Survey

In total, N=44 eligible participants completed the online survey over 3 months (Table 3).

**Table 3.** Nature of online survey participants

| Stakeholder type | n |
| --- | --- |
| Prison health provider | 15 |
| Corrective services (custody and community) | 14 |
| Post-release transition support services | 5 |
| Advocacy and community groups | 2 |
| Researcher | 5 |
| Other government agency | 2 |

#### Importance and level of agreement

All respondents (N=44) ranked the 37 recommendations in terms of importance. The results are displayed in Table 4. Each item is presented in order of their median importance score and level of agreement (calculated as the mean absolute deviation). All but one of the recommendations were strongly supported, indicated by a median score of 7-9.

**Table 4.**
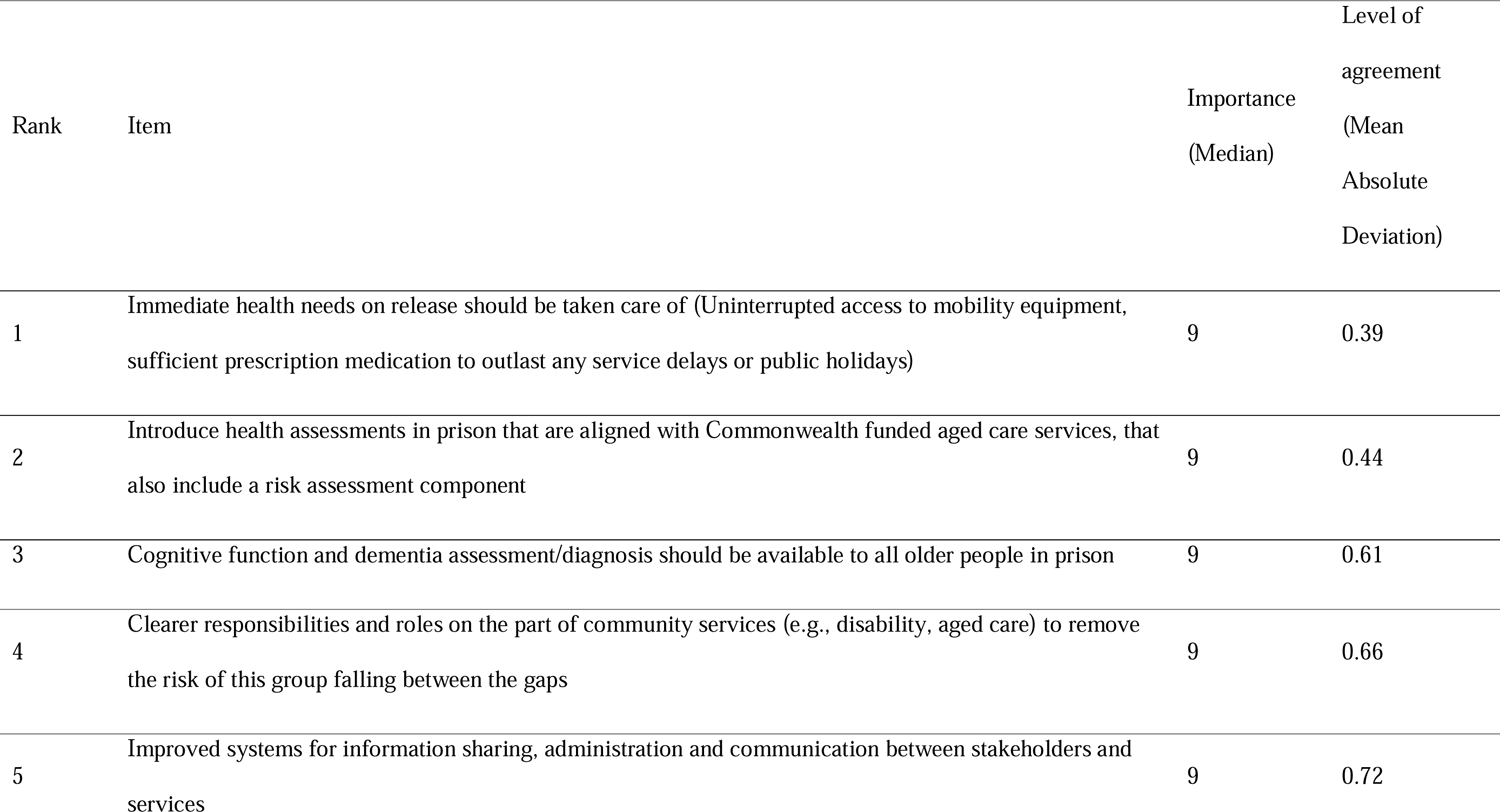
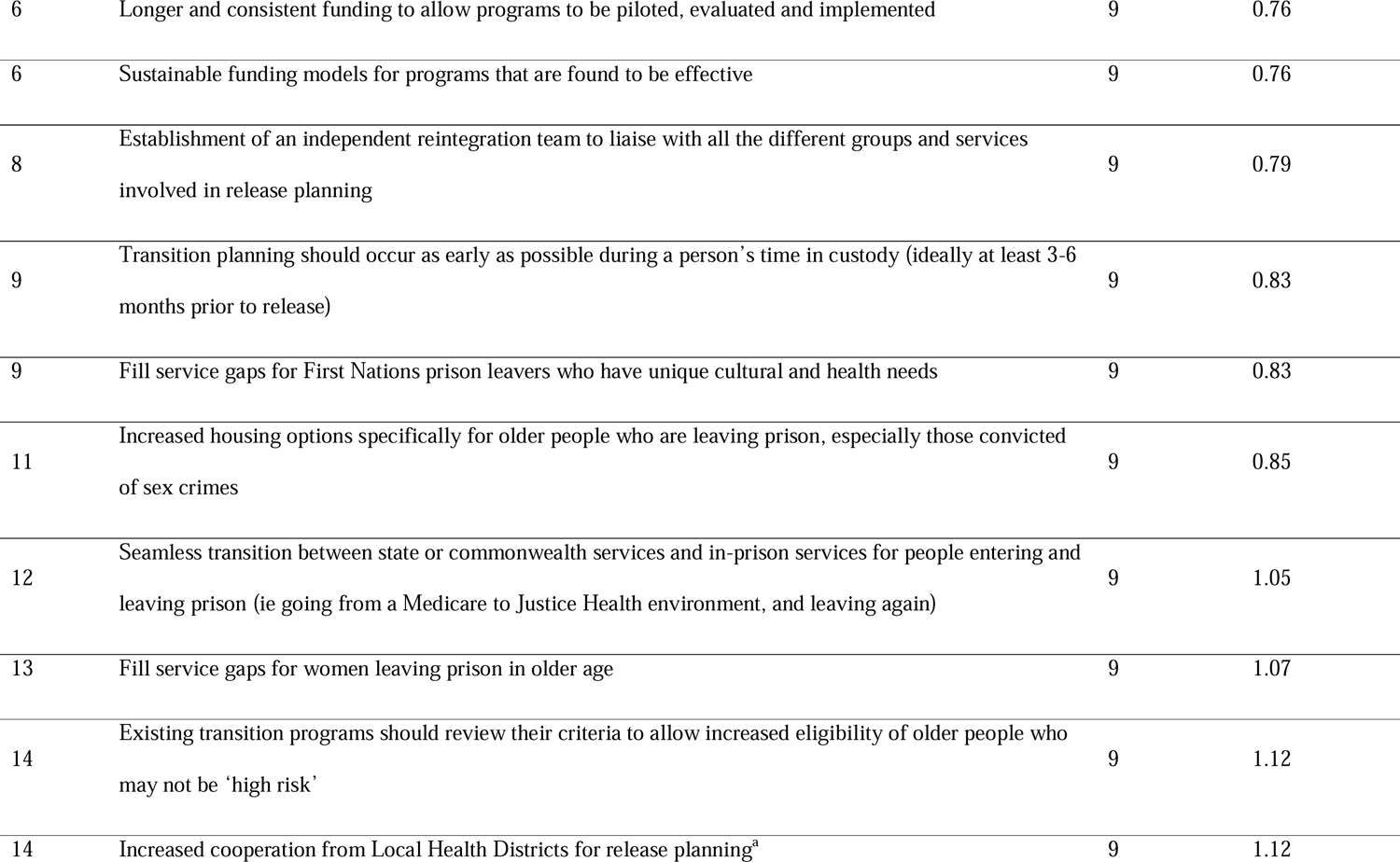
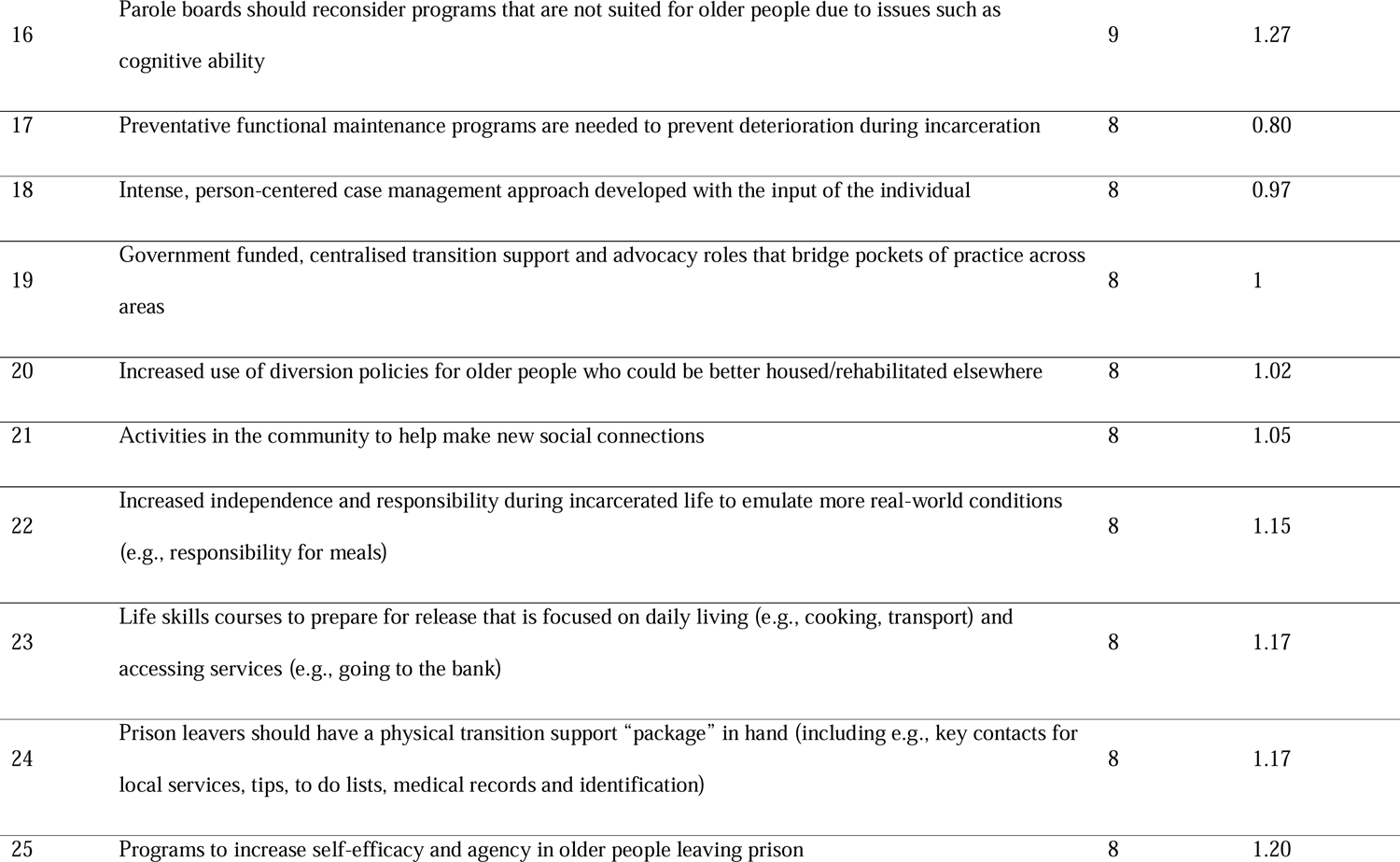
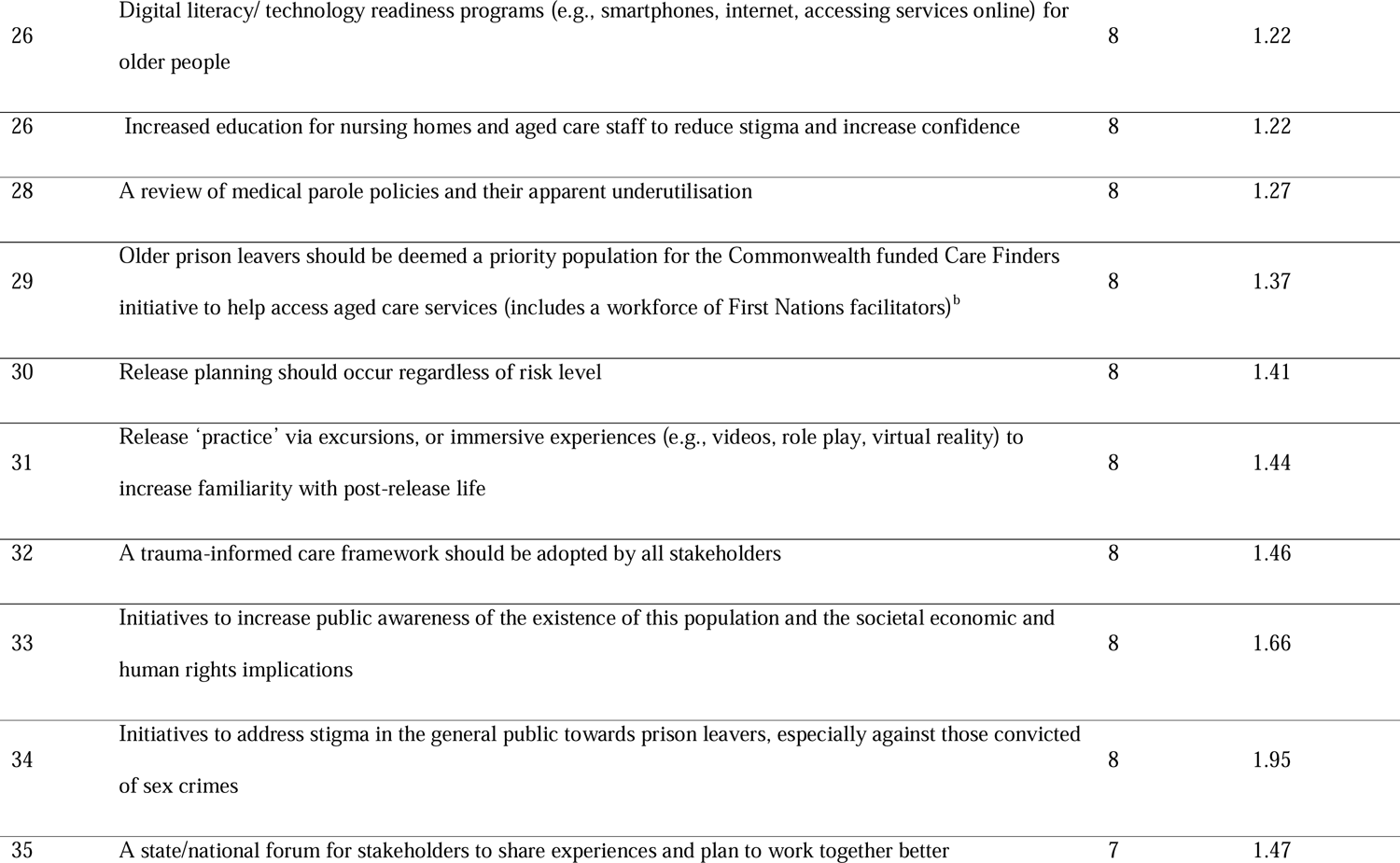
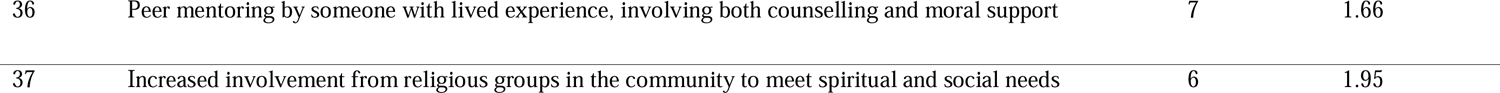
Final recommendations in order of importance (median score) and level of agreement (mean absolute deviation)

#### Priorities for implementation by stakeholder group

After ranking the importance of each recommendation, participants chose the Top 7 recommendations that they believed were priorities for implementation (Table 5). The priority ranks and total score for each recommendation are presented first for the entire sample (N=44), followed by a breakdown by stakeholder type. Groups with small sample sizes (N<5; transition support workers, advocacy groups and other government stakeholders) were collapsed into one. The top priority was common across all groups except for the researcher group. Whilst there was some spread evident in the top priorities across stakeholder groups, there was a general tendency for the higher-ranking priorities to be common across groups. For example, the top 4 priorities for each group all appear within the first 9 lines of Table 5.

**Table 5.**
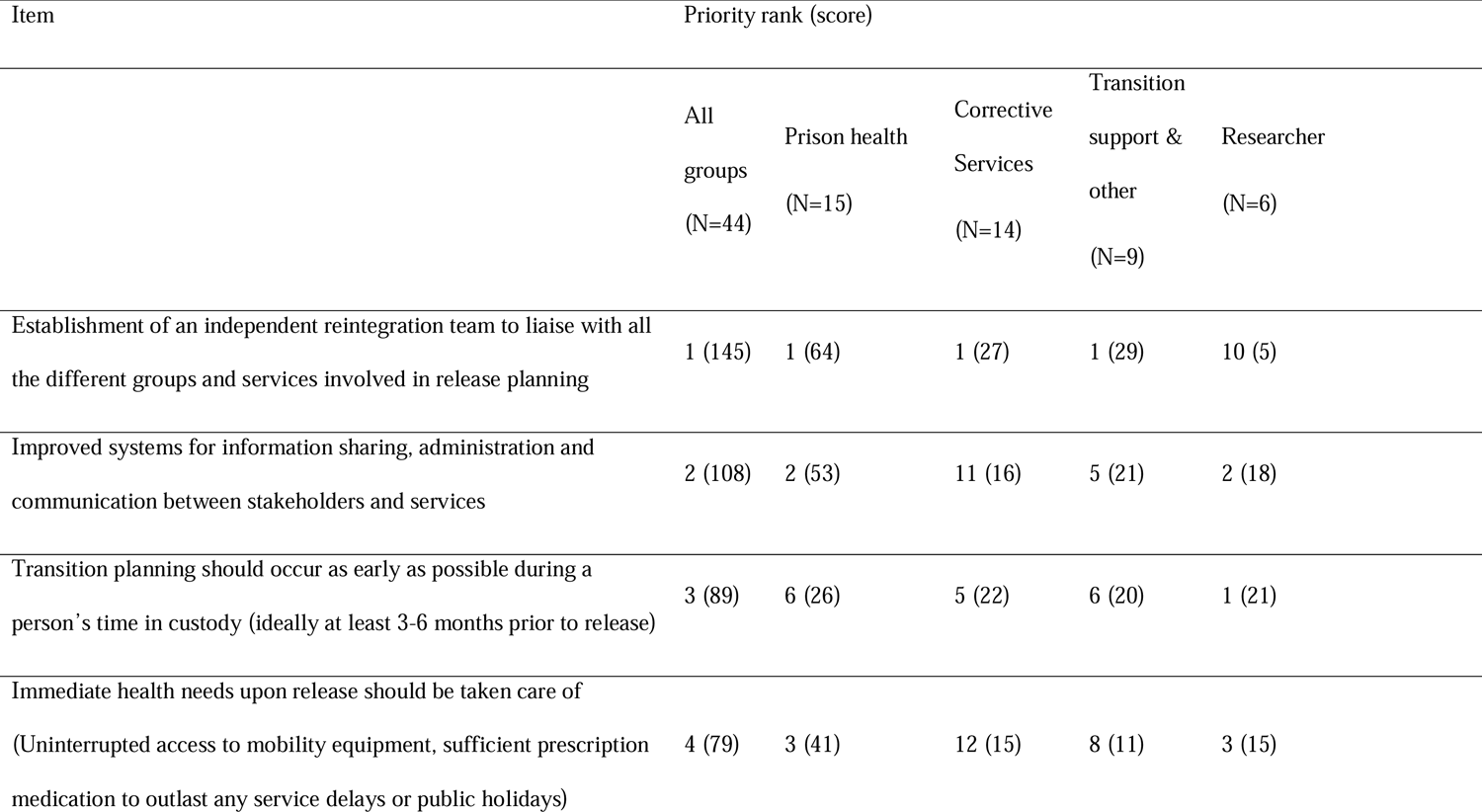
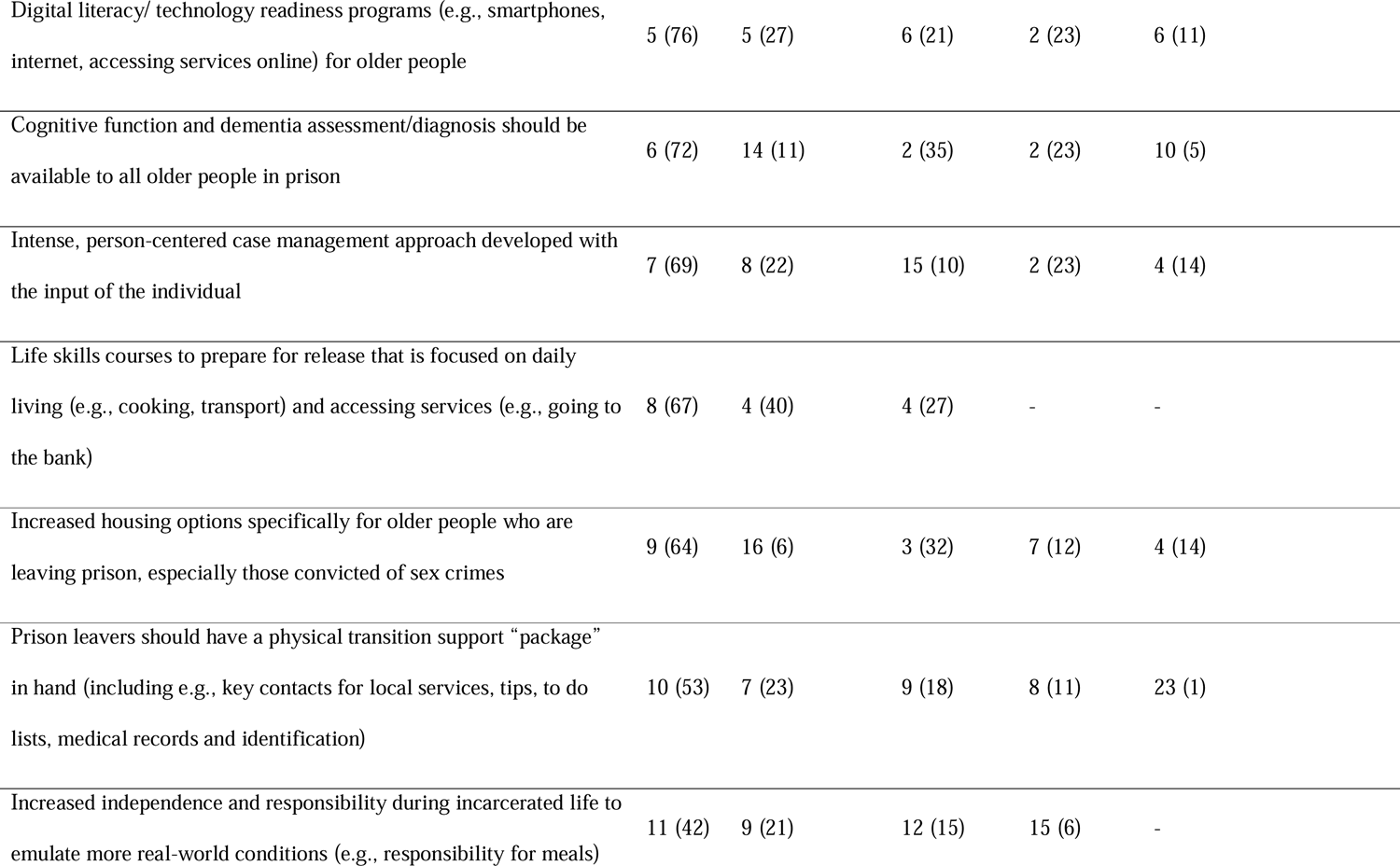
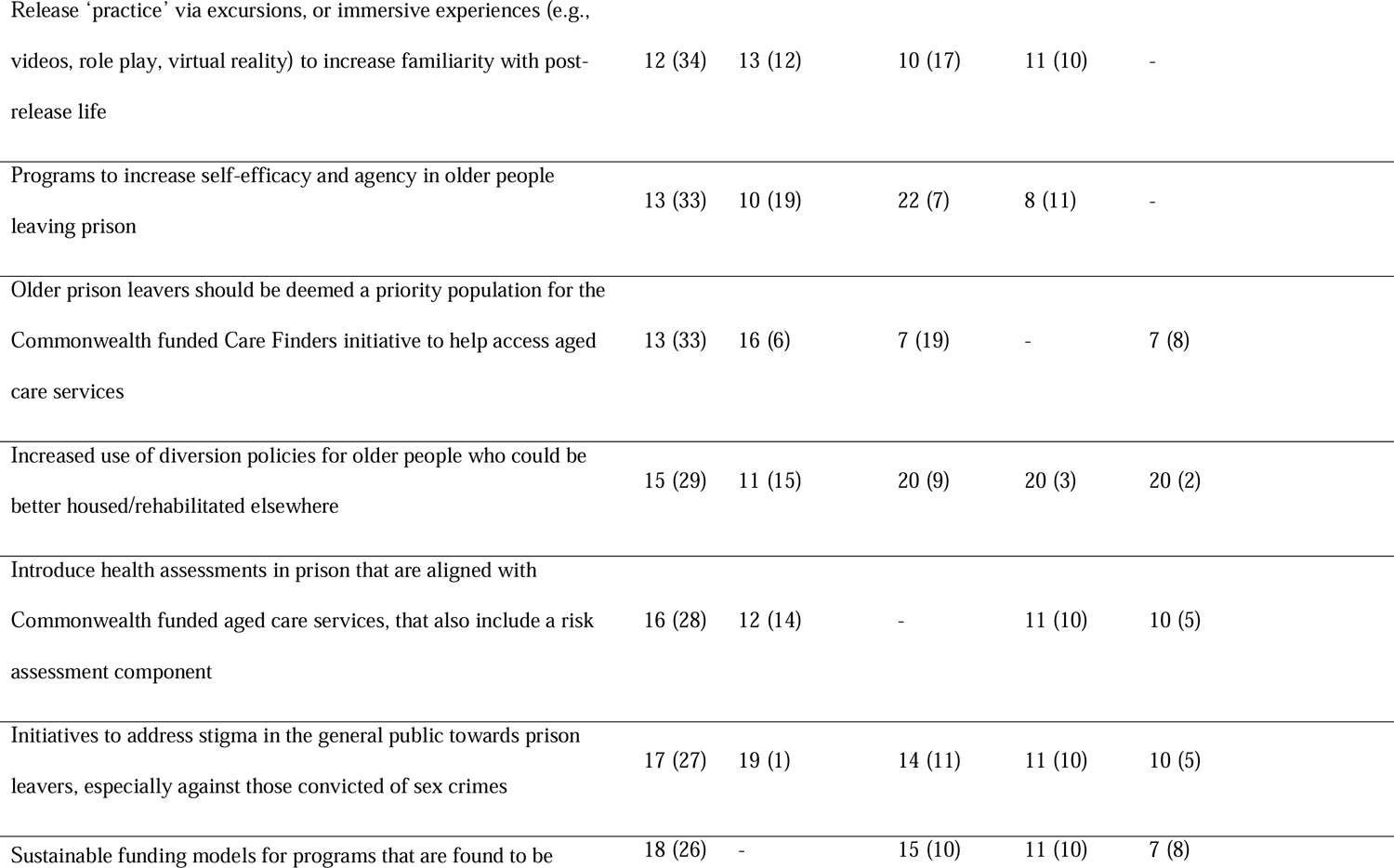
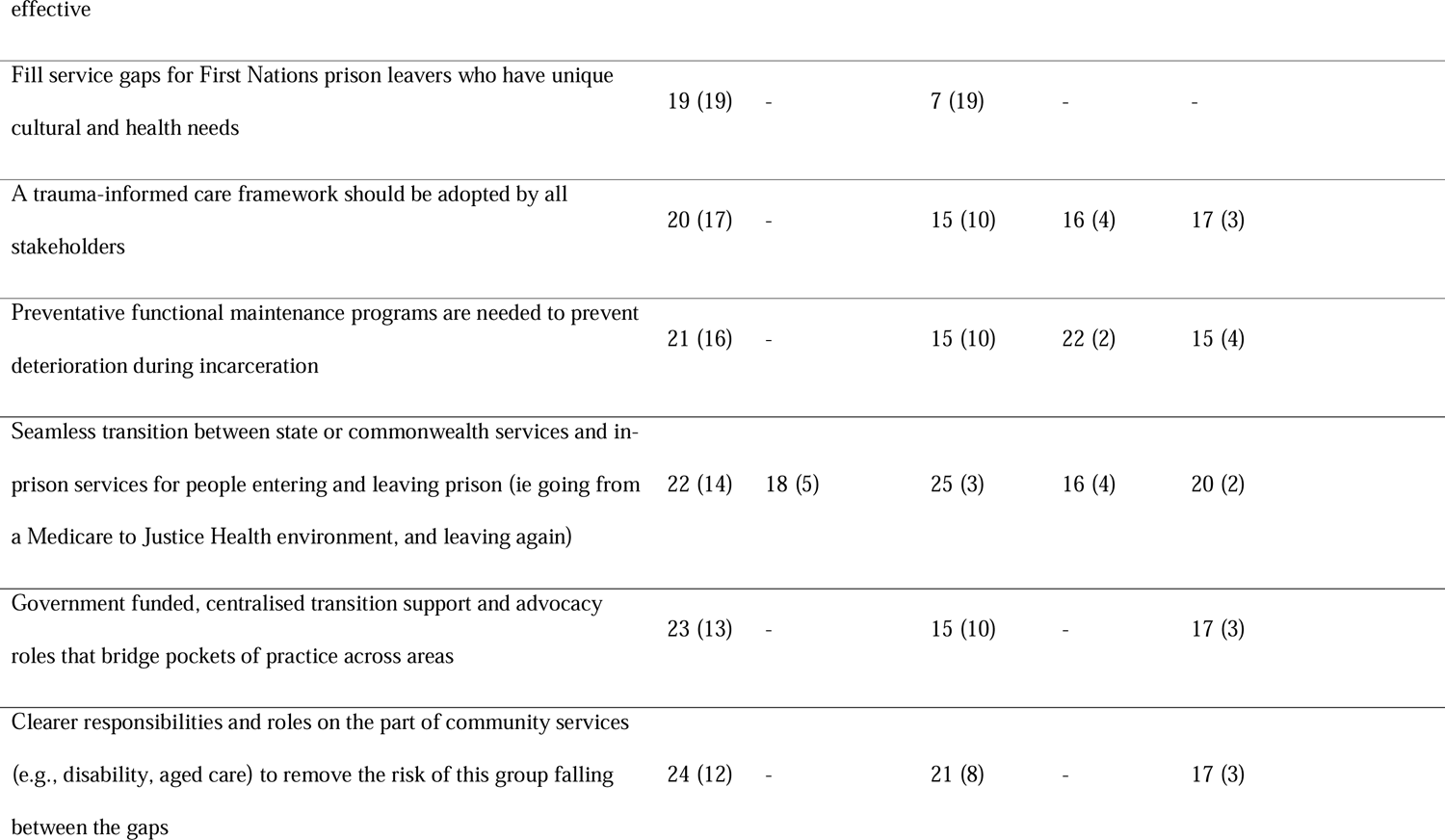
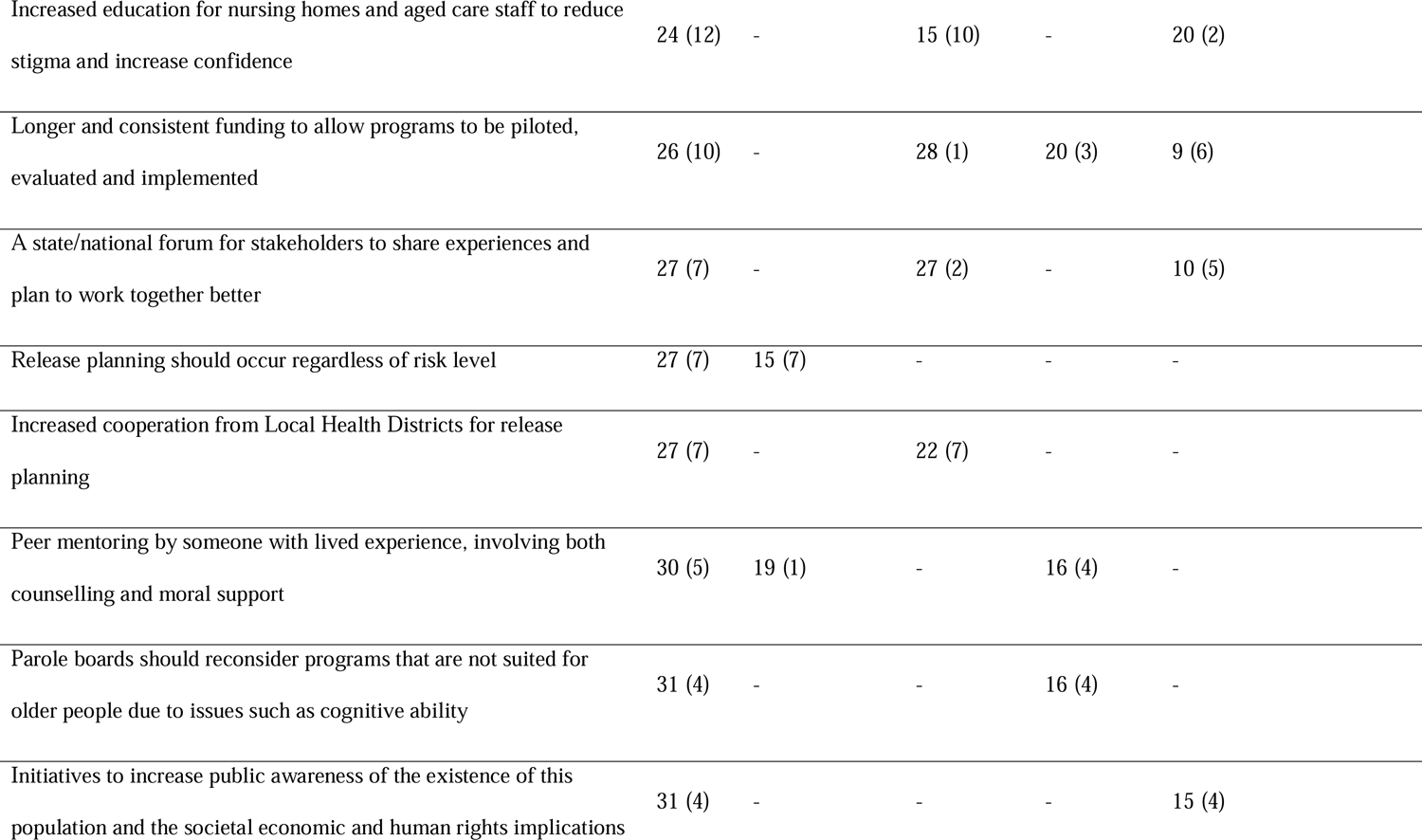
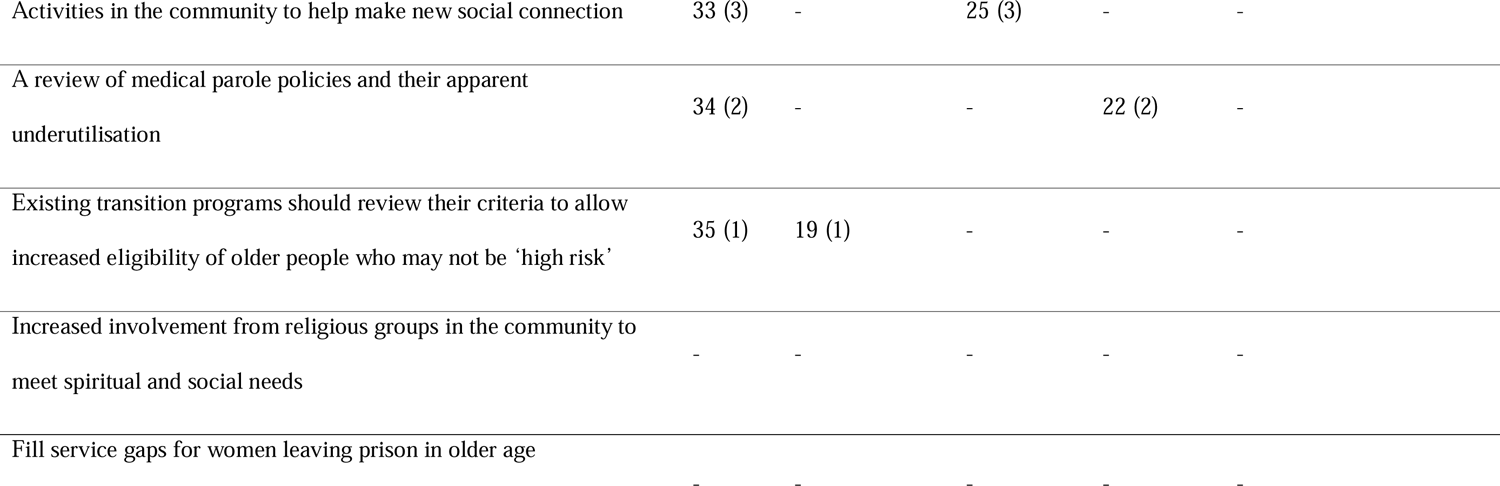
Priority rank and score for all recommendations by stakeholder type

| Item | Priority rank (score) |  |  |  |  |
| --- | --- | --- | --- | --- | --- |
|  | All groups<br>(N=44) | Prison health<br>(N=15) | Corrective Services<br>(N=14) | Transition support & other<br>(N=9) | Researcher<br>(N=6) |
| Establishment of an independent reintegration team to liaise with all the different groups and services involved in release planning | 1 (145) | 1 (64) | 1 (27) | 1 (29) | 10 (5) |
| Improved systems for information sharing, administration and communication between stakeholders and services | 2 (108) | 2 (53) | 11 (16) | 5 (21) | 2 (18) |
| Transition planning should occur as early as possible during a person's time in custody (ideally at least 3-6 months prior to release) | 3 (89) | 6 (26) | 5 (22) | 6 (20) | 1 (21) |
| Immediate health needs upon release should be taken care of<br>(Uninterrupted access to mobility equipment, sufficient prescription medication to outlast any service delays or public holidays) | 4 (79) | 3 (41) | 12 (15) | 8 (11) | 3 (15) |
| Digital literacy/ technology readiness programs (e.g., smartphones, internet, accessing services online) for older people | 5 (76) | 5 (27) | 6 (21) | 2 (23) | 6 (11) |
| Cognitive function and dementia assessment/diagnosis should be available to all older people in prison | 6 (72) | 14 (11) | 2 (35) | 2 (23) | 10 (5) |
| Intense, person-centered case management approach developed with the input of the individual | 7 (69) | 8 (22) | 15 (10) | 2 (23) | 4 (14) |
| Life skills courses to prepare for release that is focused on daily living (e.g., cooking, transport) and accessing services (e.g., going to the bank) | 8 (67) | 4 (40) | 4 (27) | - | - |
| Increased housing options specifically for older people who are leaving prison, especially those convicted of sex crimes | 9 (64) | 16 (6) | 3 (32) | 7 (12) | 4 (14) |
| Prison leavers should have a physical transition support “package” in hand (including e.g., key contacts for local services, tips, to do lists, medical records and identification) | 10 (53) | 7 (23) | 9 (18) | 8 (11) | 23 (1) |
| Increased independence and responsibility during incarcerated life to emulate more real-world conditions (e.g., responsibility for meals) | 11 (42) | 9 (21) | 12 (15) | 15 (6) | - |
| Release 'practice' via excursions, or immersive experiences (e.g., videos, role play, virtual reality) to increase familiarity with post-release life | 12 (34) | 13 (12) | 10 (17) | 11 (10) | - |
| Programs to increase self-efficacy and agency in older people leaving prison | 13 (33) | 10 (19) | 22 (7) | 8 (11) | - |
| Older prison leavers should be deemed a priority population for the Commonwealth funded Care Finders initiative to help access aged care services | 13 (33) | 16 (6) | 7 (19) | - | 7 (8) |
| Increased use of diversion policies for older people who could be better housed/rehabilitated elsewhere | 15 (29) | 11 (15) | 20 (9) | 20 (3) | 20 (2) |
| Introduce health assessments in prison that are aligned with Commonwealth funded aged care services, that also include a risk assessment component | 16 (28) | 12 (14) | - | 11 (10) | 10 (5) |
| Initiatives to address stigma in the general public towards prison leavers, especially against those convicted of sex crimes | 17 (27) | 19 (1) | 14 (11) | 11 (10) | 10 (5) |
| Sustainable funding models for programs that are found to be | 18 (26) | - | 15 (10) | 11 (10) | 7 (8) |
| Fill service gaps for First Nations prison leavers who have unique cultural and health needs | 19 (19) | - | 7 (19) | - | - |
| A trauma-informed care framework should be adopted by all stakeholders | 20 (17) | - | 15 (10) | 16 (4) | 17 (3) |
| Preventative functional maintenance programs are needed to prevent deterioration during incarceration | 21 (16) | - | 15 (10) | 22 (2) | 15 (4) |
| Seamless transition between state or commonwealth services and in-prison services for people entering and leaving prison (ie going from a Medicare to Justice Health environment, and leaving again) | 22 (14) | 18 (5) | 25 (3) | 16 (4) | 20 (2) |
| Government funded, centralised transition support and advocacy roles that bridge pockets of practice across areas | 23 (13) | - | 15 (10) | - | 17 (3) |
| Clearer responsibilities and roles on the part of community services (e.g., disability, aged care) to remove the risk of this group falling between the gaps | 24 (12) | - | 21 (8) | - | 17 (3) |
| Increased education for nursing homes and aged care staff to reduce stigma and increase confidence | 24 (12) | - | 15 (10) | - | 20 (2) |
| Longer and consistent funding to allow programs to be piloted, evaluated and implemented | 26 (10) | - | 28 (1) | 20 (3) | 9 (6) |
| A state/national forum for stakeholders to share experiences and plan to work together better | 27 (7) | - | 27 (2) | - | 10 (5) |
| Release planning should occur regardless of risk level | 27 (7) | 15 (7) | - | - | - |
| Increased cooperation from Local Health Districts for release planning | 27 (7) | - | 22 (7) | - | - |
| Peer mentoring by someone with lived experience, involving both counselling and moral support | 30 (5) | 19 (1) | - | 16 (4) | - |
| Parole boards should reconsider programs that are not suited for older people due to issues such as cognitive ability | 31 (4) | - | - | 16 (4) | - |
| Initiatives to increase public awareness of the existence of this population and the societal economic and human rights implications | 31 (4) | - | - | - | 15 (4) |
| Activities in the community to help make new social connection | 33 (3) | - | 25 (3) | - | - |
| A review of medical parole policies and their apparent underutilisation | 34 (2) | - | - | 22 (2) | - |
| Existing transition programs should review their criteria to allow increased eligibility of older people who may not be 'high risk' | 35 (1) | 19 (1) | - | - | - |
| Increased involvement from religious groups in the community to meet spiritual and social needs | - | - | - | - | - |
| Fill service gaps for women leaving prison in older age | - | - | - | - | - |

## Discussion

### Overview of findings

This deliberative study produced 36 recommendations for improving the health and social outcomes of older people leaving prison with strong support from multiple stakeholders, and one recommendation with moderate support. Overall, the underlying issues were aligned with those identified in other countries such as the United States and United Kingdom (Metzger et al., 2017; O’Hara et al., 2015), with some more specific suggestions relevant to the Australian context. The findings confirm the recommendations made in a previous Australian roundtable on supporting people released from prison (i.e., referral practices, health needs, housing and funding stability) (Schwartz et al., 2020), whilst extending these to consider multistakeholder cooperation and issues that are pertinent to this age group. Overall, the study findings may be synthesised into two important lessons regarding the growth of this population, that are discussed further in proceeding sections:

1. They are a high-needs, unique, and underserved group at risk of significant health and social inequity in the community
2. Multi-sector stakeholder cooperation will be crucial to service this growing group

The ranking of priorities for implementation should be interpreted with care due to the small number of respondents in certain stakeholder groups. All but one of the recommendations were voted with strong support. Thus, the overall order of the priorities in Table 5 should not be interpreted as an indication of which recommendations are more important than others.

Rather, the order reflects a combination of urgency for implementation, as well the likelihood of multistakeholder support. The differences in ranking between stakeholder groups reflects that each group has experienced varied exposure to different parts and consequences of the release process and may indicate which groups may be poised to provide more (or less) support or drive certain recommendations.

### Key issues and implications

#### High-needs, unique and underserved group

These findings identify that older people leaving prison have high and complex needs that are not well serviced with existing practices and resources. Whilst the areas of concern are similar to that of younger people leaving prison, there are unique and escalated needs for this age group, which stem from: more complex and heightened health needs at release, higher risk of homelessness, often longer experiences of institutionalisation, lack of age-appropriate care and release planning whilst incarcerated, and a lack of advocacy. Five of the recommendations thus call for changes in practices for services dealing with this population during and after release (Appendix 1, Category E). These suggestions touch on three specific areas including trauma-informed care, using ‘needs’ rather than ‘risk’ as the determining factor for service allocation in this group, and alternative housing (outside of prison in other secure settings) for those who are unwell and a low risk to the community.

A repeated concern is the risk of this population falling through service gaps in the community. According to these findings, these gaps emerge because of the multifaceted nature of the older person’s needs, the lack of support and advocacy to bridge these needs, a lack of continuity between in prison and community services, and unclear roles on the part of each ‘receiving’ service in the community (e.g., aged care, disability services) in meeting these needs. Older women and people who are Aboriginal or Torres Strait Islander were identified as particularly vulnerable. This is in line with existing literature that recognises these groups experience additional health needs that intersect with old age and imprisonment, that often remain unmet after release (Abbott et al., 2017; Day & Tamatea, 2019; Handtke et al., 2015).

Also evident is apparent inadequacy in release planning for this age group. Stakeholders supported the need for individuals to access a broader range of health assessments and support resources to take care of their health and service needs after release. This aligns with evidence indicating that this group has high and immediate needs after release due to multiple health conditions, lost social connections, housing and financial instability (Australian Institute of Health and Welfare, 2019; Lares & Montgomery, 2020), as well as international studies indicating that release planning for this age group is both lacking and entails significant negative consequences (Forsyth et al., 2015; Hagos, 2021; O’Hara et al., 2015).

Importantly, additional resources are needed to adequately service this growing group. Two of the recommendations (including the top-ranked priority) called for independent, government-funded roles or services to liaise between the multiple stakeholder groups, and to provide advocacy for their needs. This reflects several underlying issues. That is, the needs of older prison leavers span multiple services and sectors with significant work involved in liaising with each of these. Also, these tasks do not naturally fit within the remit of any existing staff roles whether in health, community services, or correctional services. Finally, existing staff are not adequately resourced to complete these tasks. Thus, there is a need for an independent and dedicated role or service to be created.

This ambiguity of staff roles arising from the complex, interconnected health, social and criminogenic needs of older people leaving prison has been raised in previous research (Hagos et al., 2021). The preference for an independent role may reflect a lack of resources on the part of existing staff to complete these tasks, and/or the belief that no existing service is currently suitable to take on this role. In either case, with the rise in this population a new area of need has arisen that requires resources to adequately service. A review of the additional tasks arising at the intersection of these multiple services, and the unique expertise that different stakeholders may already have, will provide direction on how to begin servicing this need. Economic evaluations of the potential benefits of such a role in improving public health outcomes, reducing unemployment and recidivism, can assist in this case.

An important hurdle identified in this work for servicing needs and allocation of resources, is the stigma experienced from services and people in the community. The recommendations call for increased advocacy, awareness and reduced stigma, particularly in securing aged care and housing. Seeking stable housing is a challenge for people leaving prison at all ages (Schwartz et al., 2020). Additional complexity is present where aged care is needed in an already-limited housing landscape. Advocacy and stigma-reducing initiatives are particularly relevant for older people leaving prison, as they are more likely to have been imprisoned for more serious offences that carry long sentences. Discussions around this recommendation pointed to the existence of both formal (practice) and attitudinal barriers that aged care providers and the general community in allocating public resources to previously incarcerated people. The precise nature of such barriers should be explored further in qualitative work and policy reviews and followed by appropriate responses to address these.

#### The need for multistakeholder cooperation

Many highly ranked recommendations confirm the need for improved ways of working together for the multiple stakeholders involved in reintegration of older people leaving prison. This is aligned with the purpose of the COJENT framework (Metzger et al., 2017) which underlies this work, and confirms existing findings regarding disparate and siloed ways of working even between stakeholders within the prison context, such as prison health services and corrective services (e.g., Hagos, 2021). This also aligns with the World Health Organization (WHO) Prison Health Framework Priority to “Conduct intersectoral work for better performance and outcomes”(World Health Organization, 2021, p. 6).

The need for multistakeholder cooperation is illustrated well in Figure 2, where we mapped each recommendation to each stakeholder that will be primarily involved in implementing that recommendation. The volume of each chord connecting stakeholders provides an indication of how many recommendations they have in common. Inevitably, the bulk of the work will need to occur with the cooperation of corrective services (both correctional facilities and community corrections). This work urges the establishment of new or closer collaborations between corrective services and many other stakeholders in the community, with long term benefits for all stakeholders involved both in terms of resource use and more effective servicing of the health and social needs of this population.

**Figure 2.**
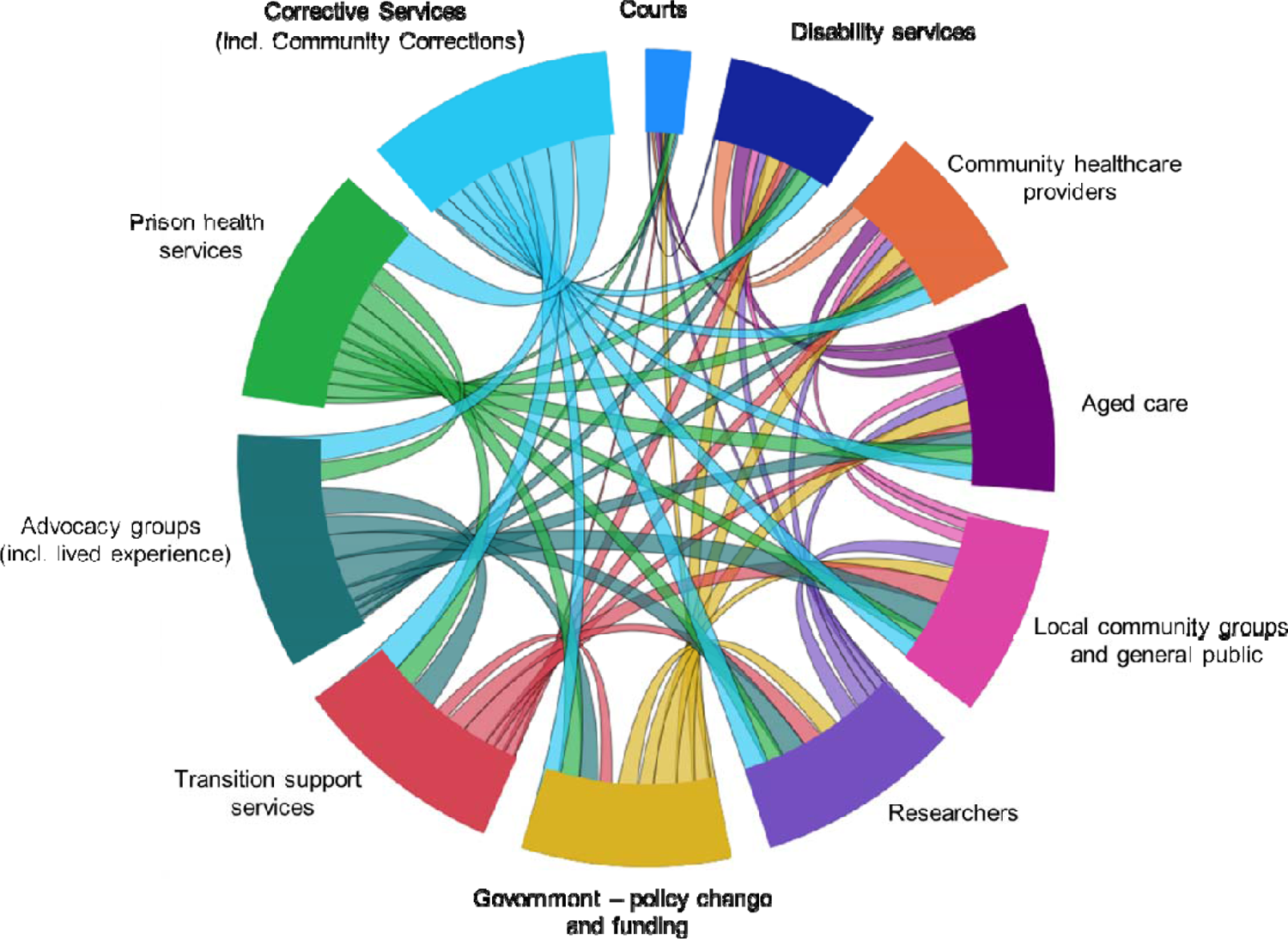
Chord diagram representing shared delivery of recommendations produced in this study. Larger chord volume indicates a higher number of shared recommendations with other stakeholders.

Throughout the recommendations there is a clear emphasis on the importance of preparing an older person for post-prison life whilst they are still in custody (Appendix 1; Categories B-D). Psychological and practical buffers are needed to ease transition between a highly regimented and uniquely restricted life in custody, and independent life in the community. Such programs are unlikely to be feasibly implemented through the expertise and resources of corrective services alone. Rather, ‘lowering the walls’ to permit older people in prison to access varying programs offered by community-based organisations prior to their release will be beneficial. In many ways this population are among the most marginalised sectors of society and have been identified as a primary target for public health interventions (Kinner & Wang, 2014; The Lancet, 2021). Existing programs for older people in the community in key areas such as health literacy and digital literacy can be brought to this population, to provide benefits not only for community reintegration but for public health and social welfare overall.

These findings show that stakeholders strongly support initiatives that work towards the ideal of reduced gaps between prison and community services. The key to achieving this, whilst also addressing the risk of falling between service gaps, appears to be in encouraging continuity of care via integrated service delivery across prison and the community. Unequal and discontinued care coverage both during incarceration and after release is a global issue (Winkelman et al., 2022), and the current findings arise in the context of sustained efforts by Australian stakeholders to lobby for the extension of the universal healthcare insurance scheme (Medicare) to people in prison with clear human rights, economic and public health benefits (Cumming et al., 2018). To our knowledge, no models of integrated service delivery between prison, health and community services have been developed or tested. Such efforts align with the principle of ‘equivalence of care’, an obligation for prison health services in Australia as well as ‘universal health coverage’ an identified priority by the World Health Organization and mandated by the UN Nelson Mandela Rules (Jotterand & Wangmo, 2014; World Health Organization, 2021, p. 6).

A strongly supported recommendation was for shared information, communication and administrative systems that are accessible across multiple organisations. This will be the capstone recommendation for enabling better working relationships among stakeholders. It also aligns with the WHO Prison Health Framework Priority to “Strengthen prison information systems to enhance surveillance and response capacity” (World Health Organization, 2021, p. 6), with outdated information systems identified as an ‘avoidable barrier’ to improved health outcomes in those who experience incarceration (Winkelman et al., 2022). The creation of shared systems across health, social service and correctional services across both prisons and the community is a complex task, involving cooperation between research, government and industry to ensure the solution is evidence-based and effective. Some foreseeable challenges will include differing levels of digital readiness at both systems and workforce levels, reconciling competing interests, addressing privacy or security concerns, and involving end users in a meaningful way to the design process. However, there is a clear need for a solution to address the obstacles experienced by stakeholders in accessing important health and personal information in a timely fashion.

### Strengths and Limitations

A strength of this study was in bringing together the perspectives of multiple stakeholders both in prisons and the community, to advance an agenda for policy, practice and research. To our knowledge it is the first study that has done this in Australia. In the final sample (N=44), there was strong representation from the prison health and corrective services stakeholder groups, albeit with less representation from those in research, advocacy, community services and aged care. This was due to a comparatively smaller pool of individuals with the level of expertise required to participate. In particular, only a small proportion of the sample represented aged care, advocacy and research. Despite this, aged care and advocacy issues were well-represented in the final recommendations. Overall, we believe the expertise of the selected participants in the workshops was strong, and a crucial mass of expertise on this population was achieved. Notwithstanding these limitations, the workshops that informed the recommendations were comprehensive, and resultant issues were all subsequently strongly endorsed by the wider sample. In moving forward, active involvement from appropriate stakeholders from aged care, advocacy and research in designing and implementing solutions will be important.

Due to ethical and administrative barriers and resource limitations, the study sample were predominantly those currently affiliated with two Australian jurisdictions (New South Wales and Victoria). Whilst some limit on generalisability will exist, the sample nevertheless provided strong representation of the key issues. New South Wales and Victoria represent the first and third largest population of prisoners cross the eight states and territories respectively, managing almost half (47.4%) of prisoners in Australia (ABS, 2021). Many workshop participants also (N=15) reported extensive experience and knowledge that spans multiple jurisdictions in Australia. Whilst local solutions will be needed, the arising issues appear to have universal applicability and reflect concerns in existing literature both in Australia and worldwide.

An important gap in this work is the views of those with lived experience of leaving prison in older age. Research is needed to capture both their experiences of navigating post-release life as well as their priorities and recommendations for improving outcomes during this time. This group should also be centrally involved in designing and implementing solutions.

## Conclusions

Multiple stakeholders across health, justice, research, and social services have common interests and a clear need to work together to ensure positive outcomes for the growing and vulnerable population of older people leaving prison. Resources, programs and policy change is needed at multiple levels, including the individual, organisational, governmental and wider society. There are similarities in the needs and that are documented for younger people leaving prison, and thus it is likely that implementing these recommendations will also translate to benefits for prison leavers of all ages. Whilst not all recommendations will be feasible to implement, similar solutions that meet these key areas of need should be pursued. Next steps in this work will involve the continued application of the COJENT framework to achieve this, i.e., (ii) conducting community-based needs assessments, (iii) implementing quick-response interventions, (iv) holding public forums to engage stakeholders to prepare for action (v) consolidating the evidence and engaging “champions” to collaboratively develop and deliver an action plan (Metzger et al., 2017). This work should also progress the development of a national strategy for improving release outcomes for this population, providing a practical imperative for further work.

## Declarations of interest

none

## Data availability statement

The authors do not have permission to share data.

## Data Availability

The authors do not have permission to share the data.

## Acknowledgements

The authors wish to thank the Australian Association of Gerontology Hal Kendig Research Development Program for providing financial support for the conduct of this research study. The authors also wish to acknowledge the support from the NSW Community Restorative Centre.

## Appendix 1

Category A. Working Together, Better

1. Improved systems for information sharing, administration and communication between stakeholders and services
2. Seamless transition between state or commonwealth services and in-prison services for people entering and leaving prison (ie going from a Medicare to Justice Health environment, and leaving again)
3. Clearer responsibilities and roles on the part of community services (e.g., disability, aged care) to remove the risk of this group falling between the gaps
4. Increased cooperation from Local Health Districts for release planning
5. A state/national forum for stakeholders to share experiences and plan to work together better
6. Establishment of an independent reintegration team to liaise with all the different groups and services involved in release planning
7. Government funded, centralised transition support and advocacy roles that bridge pockets of practice across areas Category B. Age-appropriate Care in Prison
8. Cognitive function and dementia assessment/diagnosis should be available to all older people in prison
9. Preventative functional maintenance programs are needed to prevent deterioration during incarceration
10. Introduce health assessments in prison that are aligned with Commonwealth funded aged care services, that also include a risk assessment component Category C. Preparing the Individual
11. Life skills courses to prepare for release that is focused on daily living (e.g., cooking, transport) and accessing services (e.g., going to the bank)
12. Digital literacy/ technology readiness programs (e.g., smartphones, internet, accessing services online) for older people
13. Increased independence and responsibility during incarcerated life to emulate more real-world conditions (e.g., responsibility for meals)
14. Programs to increase self-efficacy and agency in older people leaving prison
15. Release ‘practice’ via excursions, or immersive experiences (e.g., videos, role play, virtual reality) to increase familiarity with post-release life Category D. Intense, Attentive and Individualised Release Planning
16. Transition planning should occur as early as possible during a person’s time in custody (ideally at least 3-6 months prior to release)
17. Intense, person-centered case management approach developed with the input of the individual
18. Prison leavers should have a physical transition support “package” in hand (including e.g., key contacts for local services, tips, to do lists, medical records and identification)
19. Immediate health needs upon release should be taken care of (Uninterrupted access to mobility equipment, sufficient prescription medication to outlast any service delays or public holidays) Category E. Doing Things Differently
20. A trauma-informed care framework should be adopted by all stakeholders
21. Parole boards should reconsider programs that are not suited for older people due to issues such as cognitive ability
22. Release planning should occur regardless of risk level
23. Existing transition programs should review their criteria to allow increased eligibility of older people who may not be ‘high risk’
24. Increased use of diversion policies for older people who could be better housed/rehabilitated elsewhere
25. A review of medical parole policies and their apparent underutilisation Category F. Community Programs and Support
26. Peer mentoring by someone with lived experience, involving both counselling and moral support
27. Activities in the community to help make new social connections
28. Increased involvement from religious groups in the community to meet spiritual and social needs Category G. Advocacy, Awareness and Stigma
29. Initiatives to increase public awareness of the existence of this population and the societal economic and human rights implications
30. Increased education for nursing homes and aged care staff to reduce stigma and increase confidence
31. Initiatives to address stigma in the general public towards prison leavers, especially against those convicted of sex crimes
32. Older prison leavers should be deemed a priority population for the Commonwealth funded Care Finders initiative to help access aged care services (includes a workforce of First Nations facilitators)
33. Increased housing options specifically for older people who are leaving prison, especially those convicted of sex crimes Category H. Specific Sub-populations
34. Fill service gaps for First Nations prison leavers who have unique cultural and health needs
35. Fill service gaps for women leaving prison in older age Category I. Funding
36. Longer and consistent funding to allow programs to be piloted, evaluated and implemented
37. Sustainable funding models for programs that are found to be effective

## Notes

### Competing Interest Statement

The authors have declared no competing interest.

### Funding Statement

This study was funded by the Australian Association of Gerontology Hal Kendig Research Development Program.

